# On the robustness of ethnic and socio-cultural determinants of healthcare decision-making autonomy among Hausa, Fulani, and Kanuri women in Northern Nigeria

**DOI:** 10.64898/2026.04.21.26351355

**Authors:** Abolaji Moses Ogunetimoju, Samuel Ayoola Ajeboriogbon

## Abstract

**Background:** Women’s autonomy in healthcare decision-making has become one of the most critical yet inequitably distributed determinants of health outcomes, gender equity, and sustainable development worldwide. In Northern Nigeria, the presence of ethnic and socio-cultural inequality is frequently concealed by the aggregated statistics of a region.

**Methods:** This cross-sectional secondary analysis utilized the 2024 Nigeria Demographic and Health Survey. The sample included 9,998 married women (15–49 years) identifying as Hausa, Fulani, or Kanuri in Northern Nigeria. Healthcare autonomy was categorized as husband/partner alone, respondent alone, or joint decision-making. Analysis included weighted descriptive statistics, Rao-Scott adjusted chi-square tests for residential associations, and complex sample multinomial logistic regression to identify multivariable correlates while adjusting for sampling weights, strata, and clusters.

**Results:** Mean age was 30.38 years. Most participants lacked formal education (69.6%) and resided in rural areas (72.0%). Husband-only decision-making predominated (72.6%), while 22.5% reported joint and 4.9% independent autonomy. Joint decision-making was significantly higher in urban (33.3%) than rural areas (18.3%; Adjusted F=50.892, p<0.001). In adjusted models (Reference: Kanuri), Hausa and Fulani women had substantially lower odds of joint decision-making relative to husband-only outcomes. Rural residence correlated with lower odds of both independent and joint agency. Notably, wealth status was not a significant predictor after adjustment (p > 0.05).

**Conclusions:** Ethnicity and residence are robust determinants of healthcare autonomy among women in Northern Nigeria, persisting regardless of education or wealth. This “socio-cultural paradox” suggests that economic interventions alone are insufficient. Policies must complement socioeconomic approaches with culturally responsive strategies addressing household power dynamics and entrenched social norms.

## BACKGROUND

The problem of women not having sufficient power to decide on their own healthcare has been one of the biggest international issues during the last twenty or thirty years. This issue touches on gender equality and impedes the gains in enhancing health outcomes. Women in most low income and middle-income countries tend to rely on their husbands or relatives before they can consult health care facilities. This lag may cause severe health issues, particularly in pregnancy and childbirth [1–2].

Women’s autonomy in healthcare decision-making is widely recognised as a fundamental dimension of gender equity and a critical determinant of health outcomes. In low- and middle-income countries (LMICs), restricted female agency in health matters is directly linked to delays in care-seeking, underutilisation of maternal health services, and elevated maternal and child mortality [1,2]. The World Health Organization has identified women’s empowerment as integral to achieving Sustainable Development Goal 3.1, which targets reductions in maternal mortality by 2030 [2]. Yet despite decades of global advocacy, a substantial proportion of women in LMICs continue to require spousal or familial approval before accessing healthcare, rendering their health outcomes contingent on factors beyond their own volition.

Early explanations for women’s limited healthcare autonomy centred primarily on educational attainment and economic status, with the assumption that improving these individual-level characteristics would translate directly into greater agency. While education and income remain important correlates, a growing body of evidence challenges the sufficiency of this socioeconomic framework [3,4]. Research across sub-Saharan Africa and South Asia has demonstrated that even educated and economically active women may be unable to exercise independent healthcare decisions when embedded in social structures governed by restrictive gender norms and patriarchal household power arrangements [3]. This phenomenon — in which material resources fail to translate into decision-making authority — has been described as the socio-cultural paradox of women’s empowerment, and it represents a critical theoretical and programmatic challenge for health systems seeking to improve gender equity in care-seeking [4].

Northern Nigeria illustrates this challenge with particular acuity. The region bears some of the worst maternal and reproductive health indicators in sub-Saharan Africa, yet national- and regional-level statistics frequently obscure the heterogeneity that exists within it. Northern Nigeria is home to a diverse array of ethnic communities, including the Hausa, Fulani, and Kanuri, each of which maintains distinct cultural traditions, household structures, and social norms that shape how gender relations and domestic authority are organised [5]. Treating Northern Nigeria as a socioculturally homogeneous region risks masking these ethnic variations and generating misleading conclusions about the determinants of women’s healthcare autonomy. Disaggregated, ethnicity-sensitive analysis is therefore not merely a methodological preference but a substantive necessity for understanding and addressing the drivers of women’s limited agency in this region [5].

Evidence from comparable settings underscores the inadequacy of relying on socioeconomic proxies alone to predict women’s healthcare autonomy. Studies have shown that in highly traditional environments, community-level gender norms and cultural expectations exert an influence that often supersedes the gains associated with individual educational or economic advancement [3]. A woman may possess the financial means to access healthcare independently yet remain constrained by the social expectation that her husband must sanction such decisions.

This dynamic is particularly pronounced in settings where patriarchal authority is institutionalised within household and community structures, producing a persistent gap between women’s capabilities and their realised agency — what Heise and colleagues describe as the structural embedding of gender inequality in health [3].

A related but distinct challenge is the gap between health knowledge and health action. Research has demonstrated that women who are aware of health risks and appropriate responses may nonetheless fail to act on this knowledge, not because of ignorance but because of the social consequences of acting without spousal consent [6]. Fear of conflict, stigma, or sanctions within the household can prevent women from independently seeking care even when they recognise its necessity. This knowledge-action gap is particularly problematic for time-sensitive conditions such as obstetric emergencies, where delays in decision-making translate directly into preventable mortality. Understanding what structural and normative factors sustain this gap — beyond individual-level education or awareness — is therefore essential for designing effective health interventions [6].

### Statement of the Problem

Notwithstanding the recognised importance of women’s healthcare autonomy as a determinant of health outcomes, a critical and underexplored problem persists in the Northern Nigerian context: the extent to which ethnicity and sociocultural factors independently determine women’s healthcare decision-making autonomy remains poorly understood and empirically underexamined. Existing national and regional analyses of the Nigeria Demographic and Health Survey data have tended to treat Northern Nigeria as a socioculturally uniform entity, aggregating data across ethnically and normatively distinct communities. This aggregation conceals the heterogeneity that exists within the region and generates misleading conclusions about the determinants of women’s agency. Specifically, the Hausa, Fulani, and Kanuri ethnic groups — which collectively constitute the dominant populations of Northern Nigeria — differ substantially in their cultural traditions, household structures, and gender norms, yet their distinct experiences of healthcare autonomy have not been rigorously compared using multivariable methods that adjust for confounding.

A further dimension of the problem concerns the dominant explanatory framework used in existing research. Socioeconomic variables — particularly education and household wealth — have been treated as the primary drivers of women’s autonomy, implying that economic development and educational expansion are sufficient policy levers. However, this framework fails to account for the possibility that cultural and normative forces tied to ethnic identity and residential context may exert an influence on women’s agency that persists independently of socioeconomic status. If ethnicity and residence are robust determinants of autonomy — that is, if their associations with decision-making outcomes remain significant after full adjustment for education, wealth, and age — then purely socioeconomic interventions will be insufficient, and culturally responsive, community-specific strategies will be essential. The present study addresses this problem directly, providing the first ethnic-disaggregated, complex-sample analysis of healthcare decision-making autonomy among Hausa, Fulani, and Kanuri women using the 2024 NDHS, and testing the robustness of ethnic and sociocultural determinants under rigorous multivariable control.

Despite the importance of ethnic and sociocultural context, the majority of existing studies on women’s healthcare autonomy in Nigeria either aggregate data at the national or regional level or focus exclusively on socioeconomic predictors, leaving the independent contribution of ethnic identity and residence largely unexamined after simultaneous adjustment for confounders [4,5]. This gap is significant: without understanding whether ethnicity remains a robust correlate of autonomy after controlling for education, wealth, and age, it is impossible to determine whether culturally tailored interventions are necessary or whether standard socioeconomic approaches would suffice. The present study directly addresses this gap by applying a rigorous complex sample analytical framework to the most recent nationally representative data available.

This study therefore aimed to assess the robustness of ethnicity and other sociocultural correlates of healthcare decision-making autonomy among Hausa, Fulani, and Kanuri women in Northern Nigeria, using data from the 2024 Nigeria Demographic and Health Survey (NDHS). Specifically, the study pursued three objectives: (i) to describe the distribution of healthcare decision-making autonomy across ethnic groups and residential settings; (ii) to examine the bivariate association between place of residence and autonomy status using design-based statistical methods; and (iii) to identify independent predictors of autonomy through complex sample multinomial logistic regression, with particular attention to whether ethnicity retains significance after adjustment for residence, education, wealth, and age. The concept of robustness — the persistence of associations across model specifications and covariate adjustments — is central to the study’s analytical framework and its contribution to the literature. By testing whether ethnic differentials in autonomy survive full adjustment for conventional socioeconomic confounders, the study provides evidence on whether cultural and normative factors operate independently of material conditions in shaping women’s healthcare agency. The findings are intended to inform the design of health programmes and policies that are not only socioeconomically sensitive but also culturally responsive — recognising that in contexts where gender norms powerfully constrain women’s agency, economic interventions alone are insufficient to achieve meaningful and sustained improvements in women’s healthcare autonomy in Northern Nigeria.

## METHODS

### Aim, Design, and Setting

The aim of this study was to assess the robustness of ethnicity and other sociocultural correlates of healthcare decision-making autonomy among Hausa, Fulani, and Kanuri women in Northern Nigeria. We conducted a cross-sectional secondary analysis of Demographic and Health Survey data for Nigeria obtained through the DHS Program [5]. The DHS uses a stratified cluster sampling design to obtain population relevant estimates.

### Participants and analytic sample

The analytic sample comprised 9,998 currently married women aged 15 to 49 years who identified as Hausa, Fulani, or Kanuri and resided in Northern Nigeria. The analysis used sampling weights and design variables for strata and clusters as reflected in the complex samples output, with 40 strata and 530 primary sampling units.

### Measures Outcome variable

Healthcare decision-making autonomy was derived from the DHS item that asks who usually decides on the respondent’s healthcare. The outcome was categorized into three groups:

1. Husband or partner alone (reference category)
2. Respondent alone (Self-Decision)
3. Respondent and husband or partner (Joint decision)

This categorization distinguishes independent agency from negotiated/shared agency and from exclusion from decision-making. “Husband or partner alone” was the reference outcome in regression models.

### Explanatory variables

- **Ethnicity:** Hausa, Fulani, Kanuri
- **Place of residence:** rural, urban
- **Educational attainment:** no education, primary, secondary, higher
- **Household wealth index:** poor, middle/rich Age was included as a continuous covariate. **Statistical analysis**

Analyses were conducted in SPSS version 27 using Complex Samples procedures. We produced weighted descriptive statistics for participant characteristics and autonomy categories.

We used the Chi-square test of Pearson to check the association of place of residence to status of autonomy among women. The Chi-square value is given as:

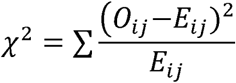

Where *o_ij_*and *E_ij_*represent observed and expected frequencies, respectively. We determined statistical significance at p <0.05.

Also, to determine the independent predictors of healthcare decision-making autonomy, we tested a multinomial logistic regression model, which is used in the case of an unordered categorical outcome. The model specification is as follows:

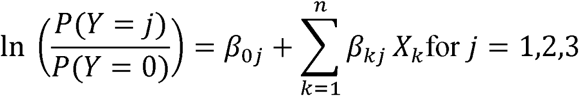

- *r*= 0 denotes husband-only decision-making (reference category),
- *j* represents alternative autonomy outcomes,
- *x_k_* are explanatory variables
- *β_kj_* are estimated parameters.

The model estimates were given in the form of Odds Ratios (ORs) with 95% Confidence Intervals (CIs). In the ethical consideration, the study used only anonymous secondary data.

### Figures

Two stacked bar charts were created: Figure 1 shows autonomy by ethnic group. Figure 2 shows autonomy by residence.

**Figure 1.**
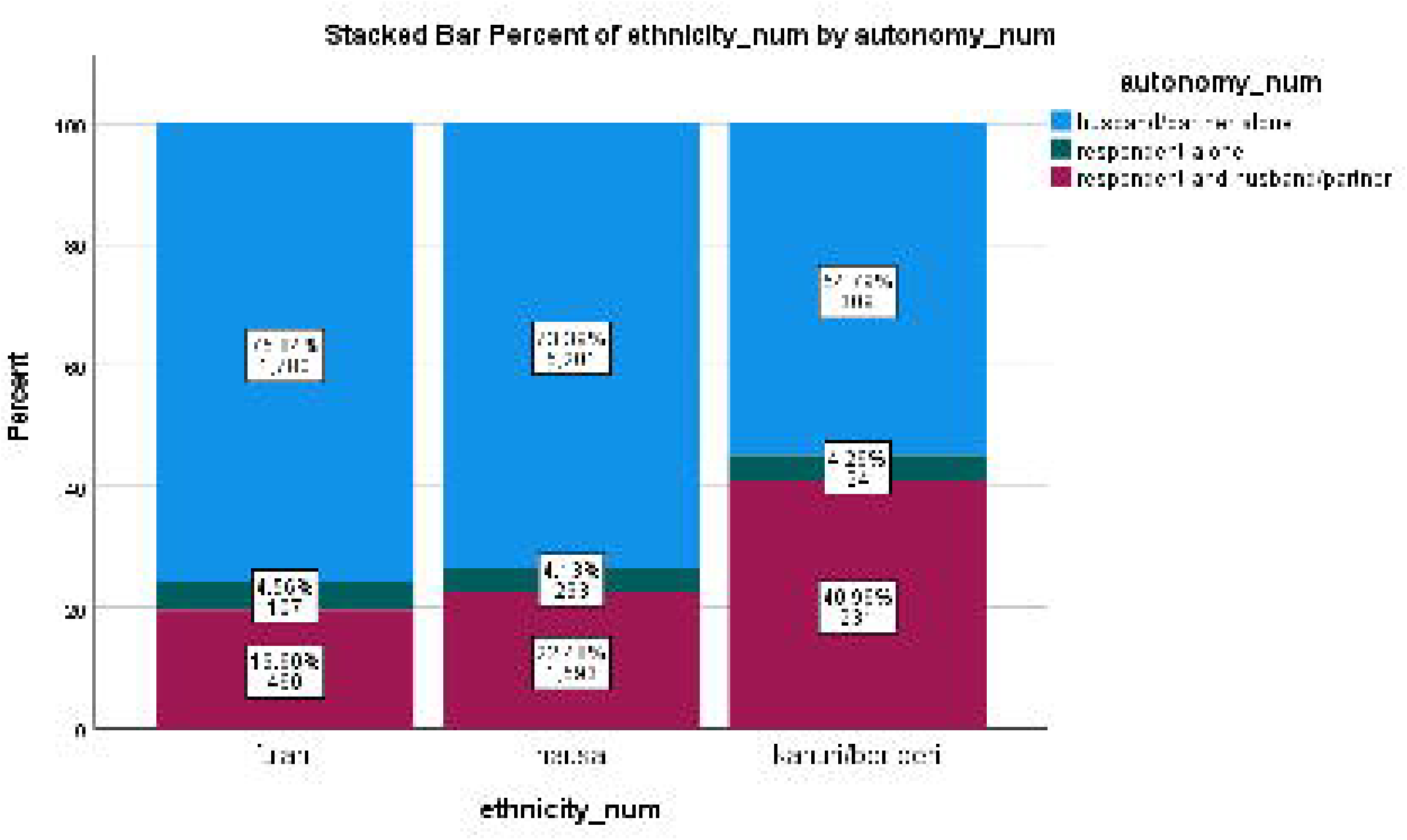
Stacked bar chart showing healthcare decision-making autonomy by ethnic group. There are two significant things that Figure 1 unveils at once. To begin with, male-dominated healthcare decision-making is a common fact among all three ethnicities - none of the groups can get away with it. Second, ethnic difference is significant, especially among the Kanuri/Beriberi women, who seem to be more collaborative when making healthcare decisions than Fulani and Hausa women. This ethnic difference is worth considering. The pattern of the Kanuri/Beriberi group may be representative of cultural norms of spousal consultation, varying household structures, or gender attitudes on a community level that do not conform to Fulan and Hausa cultures. It is interesting to add, though, that the Kanuri/Beriberi sample is much smaller (n = 564) than Hausa (n = 7,087), therefore, the result should be viewed with at least some caution.

**Figure 2.**
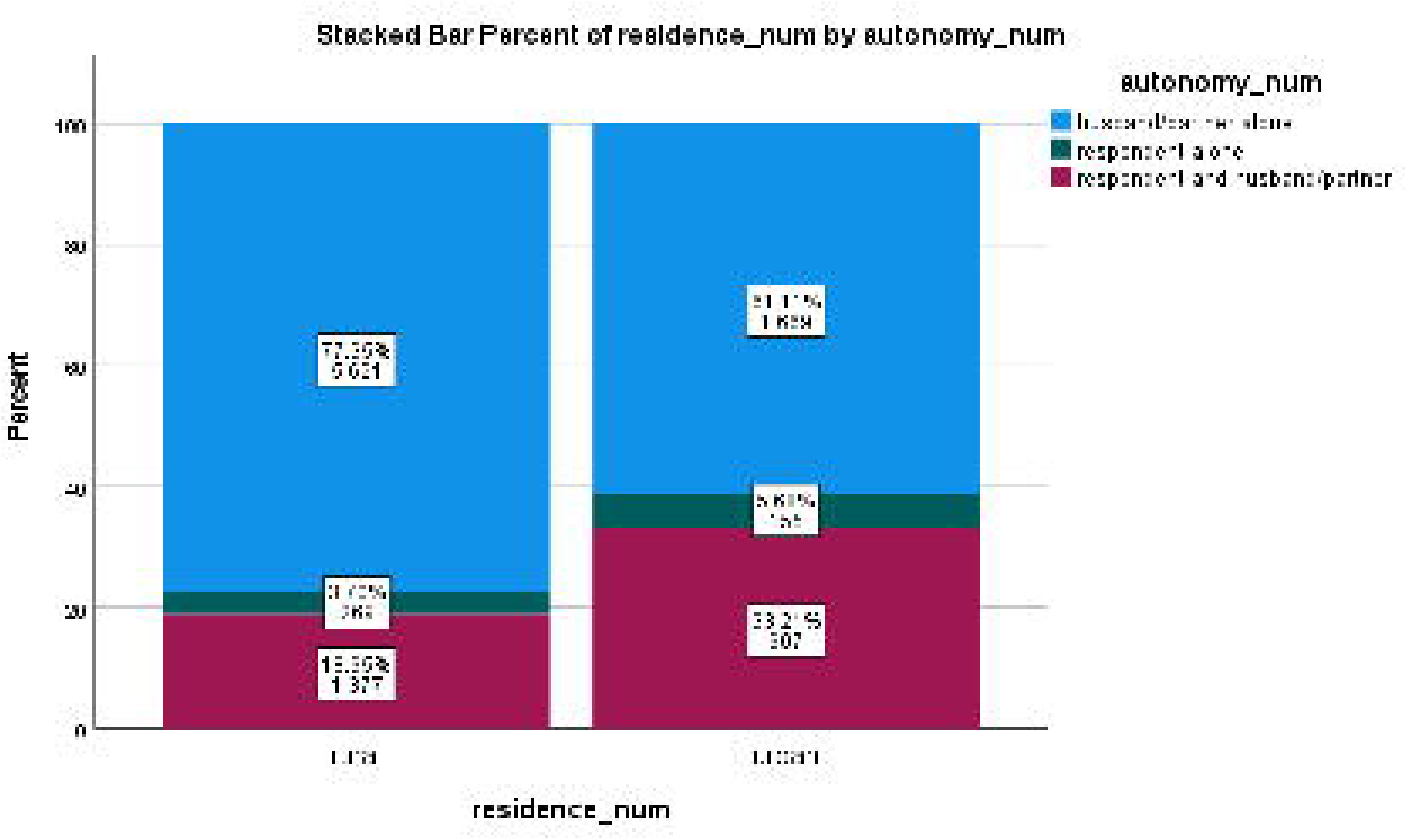
Stacked bar chart showing healthcare decision-making autonomy by residence. What Figure 2 helps us see is that no matter where they are, women have their healthcare autonomy restricted, although urban dwelling is linked to a somewhat more balanced relationship. The urban-rural difference in joint decision-making among women of about 14 percentage points indicates that urbanization could create small but significant platforms of women voice in healthcare. Nevertheless, in urban areas, most women are not the decision makers regarding their health. This is not a coincidental result and it is repeated in both groups, making it a dependable finding instead of a statistical accident, particularly with the number of women involved (more than 7,000 rural and close to 2,800 urban respondents).

### Ethics

This study used anonymized secondary data from the NDHS 2024, accessed with permission. NDHS surveys are implemented with prior ethical approval and informed consent procedures for participants.

## RESULTS

Based on a weighted analysis of **9,998** Hausa, Fulani, and Kanuri women, the data reveals a demographic facing significant structural and social barriers to healthcare. As shown in **Table 1**, this cohort demonstrates considerable socioeconomic disadvantages and highly centralized household power arrangements, where healthcare agency remains primarily under the authority of male partners. The mean age of the cohort was 30.38 years (SE = 0.15), indicating that the sample comprises women in their peak reproductive years — a period during which healthcare access and decision-making authority are particularly consequential for maternal and child health outcomes. Notably, 69.6% of participants had no formal education and 60.1% were classified as poor, representing a compounded socioeconomic vulnerability characterised by low health literacy and limited financial resources. The overwhelmingly rural composition of the sample (72.0%) further suggests that geographic barriers to healthcare access compound these socioeconomic disadvantages. The distribution of healthcare decision-making autonomy reveals a stark gender imbalance: husband or partner alone decision-making predominated at 72.6%, while joint decision-making was reported by 22.5% of women and fully independent decision-making by only 4.9%. These figures indicate that among nearly three-quarters of women in this sample, healthcare-seeking is contingent upon male consent rather than the woman’s own volition. The sociodemographic profile of this sample — dominated by Hausa women (77.6%) with high rates of low educational attainment, poverty, and rural residence — underscores that poor health outcomes in this region are not merely clinical problems but are structurally embedded in household power arrangements. Effective health system responses must therefore extend beyond service delivery to address the structural determinants of women’s agency, including the active engagement of male partners who function as primary gatekeepers of healthcare access.

**Table 1:** Sociodemographic characteristics and healthcare decision-making autonomy among Hausa, Fulani, and Kanuri women in Northern Nigeria (unweighted N 9,998)

| Characteristic | Category | Unweighted n | Weighted % | Standard error % |
| --- | --- | --- | --- | --- |
| Age | Mean (years) | 9,998 | 30.38 | 0.15 |
| Education level | No education | 7,348 | 69.6% | 1.4% |
|  | Primary | 862 | 10.2% | 0.6% |
|  | Secondary | 1,418 | 16.3% | 0.9% |
|  | Higher | 370 | 3.9% | 0.4% |
| Wealth status | Poor | 6,351 | 60.1% | 1.7% |
|  | Middle and rich | 3,647 | 39.9% | 1.7% |
| Residence | Rural | 7,267 | 72.0% | 1.4% |
|  | Urban | 2,731 | 28.0% | 1.4% |
| Ethnicity | Hausa | 7,087 | 77.6% | 1.5% |
|  | Fulani | 2,347 | 17.8% | 1.4% |
|  | Kanuri | 564 | 4.6% | 0.6% |
| Autonomy | Husband or partner alone | 7,290 | 72.6% | 0.8% |
|  | Respondent alone | 424 | 4.9% | 0.3% |
|  | Respondent and husband or partner | 2,284 | 22.5% | 0.8% |

**Source:** *Authors' analysis of DHS data using complex sample weights.*
**Legend:** Descriptive statistics for married women (15–49) in Northern Nigeria using 2024 NDHS data. Estimates are weighted to account for complex survey design.

### Autonomy patterns by ethnicity and residence

Figure 1 shows the distribution of autonomy categories by ethnic group. Figure 2 shows the same distribution by residence. Visual patterns align with multivariable results indicating lower odds of joint decision making among Hausa and Fulani women compared with Kanuri women, and lower joint decision making in rural areas compared with urban areas.

### Association between residence and autonomy

Residence was strongly associated with autonomy (adjusted F = 50.892, p < 0.001). In rural areas, 77.3% of women reported husband or partner alone decision making, compared with 60.4% in urban areas. Joint decision making was 18.3% in rural areas and 33.3% in urban areas.

Husband or partner alone decision-making was substantially more prevalent among rural women (77.3%, 95% CI: 75.5%–79.1%) than urban women (60.4%, 95% CI: 56.7%–63.9%), representing a difference of approximately 17 percentage points. Independent decision-making remained rare across both settings, recorded at 4.4% (95% CI: 3.6%–5.2%) among rural women and 6.4% (95% CI: 5.3%–7.7%) among urban women, indicating that complete female agency over healthcare is nearly non-existent regardless of residential context. The most substantively important urban-rural difference was observed in joint decision-making, which was reported by 18.3% of rural women (95% CI: 16.7%–20.0%) compared with 33.3% of urban women (95% CI: 30.0%–36.7%). Overall, table 2 establishes that rural women face a compounded deficit in healthcare autonomy, being simultaneously more likely to be excluded entirely from decision-making and less likely to participate in any shared arrangement. These findings underscore the importance of targeting rural communities as priority settings for culturally responsive interventions aimed at strengthening women’s healthcare agency in Northern Nigeria.

**Table 2:** Bivariate Analysis of Residence and Healthcare Decision-Making Autonomy.

| **Residence** | **Husband or partner alone %**<br>**(95% Confidence Interval)** | **Respondent alone %**<br>**(95% Confidence Interval)** | **Joint decision %**<br>**(95% Confidence Interval)** |
| --- | --- | --- | --- |
| Rural | 77.3% (75.5% to 79.1%) | 4.4% (3.6% to 5.2%) | 18.3% (16.7% to 20.0%) |
| Urban | 60.4% (56.7% to 63.9%) | 6.4% (5.3% to 7.7%) | 33.3% (30.0% to 36.7%) |
| **Pearson $\chi^2$** |  | **297.525** |  |
| **Adjusted F** |  | **50.892** |  |
| **P-Value** |  | **< 0.001** |  |

**Source:** Authors' analysis of DHS data using complex sample weights.
**Legend:** Design-based association using the Rao-Scott adjusted Chi-square. Degree of freedom (df1) = 1.845, (df2) = 903.845. Statistical significance was set at $p < 0.05$ .
**Interpretation:** Table 2 presents the design-based bivariate association between place of residence and healthcare decision-making autonomy among the sampled women. Residence was significantly associated with autonomy status (Rao-Scott adjusted $F = 50.892$ , $df = 1.845$ , $903.845$ , $p < 0.001$ ), confirming that where a woman lives meaningfully shapes her level of participation in healthcare decisions.

### Multivariable correlates of healthcare decision making autonomy

Taken together, Table 3 demonstrates that ethnicity and residential context are the dominant robust correlates of healthcare decision-making autonomy among Hausa, Fulani, and Kanuri women in Northern Nigeria, persisting independently of education, wealth, and age. The non-significance of wealth after adjustment, alongside the gradient pattern of education, provides compelling evidence for a socio-cultural paradox in which structural and normative factors tied to ethnic identity and geographic context exert greater influence over women’s healthcare agency than economic resources. These findings carry direct implications for the design of health interventions: programmes targeting women’s autonomy in Northern Nigeria must move beyond socioeconomic approaches and engage directly with the cultural and community-level norms that sustain male-dominated healthcare decision-making across ethnic groups and residential settings.

**Table 3.**
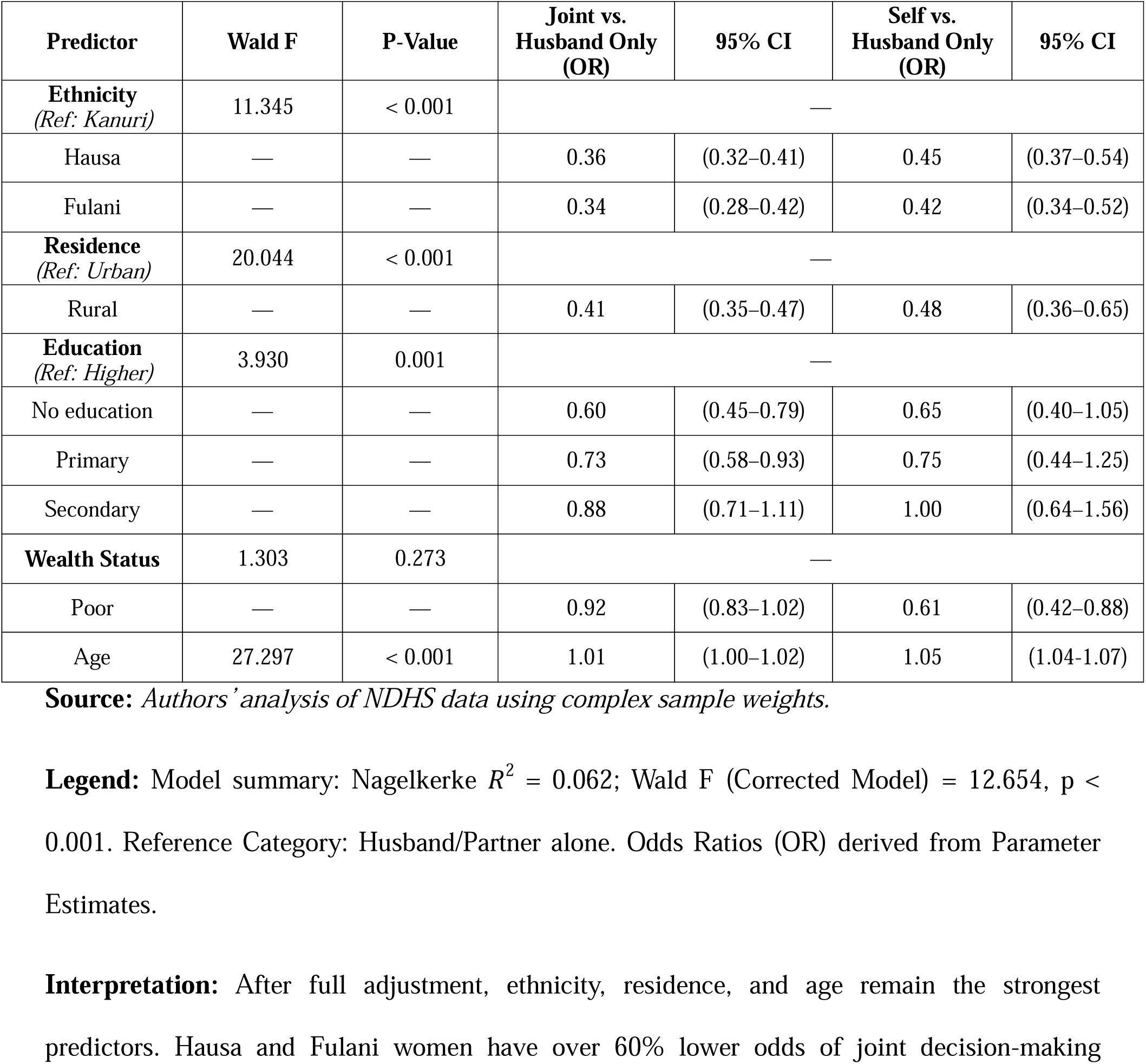
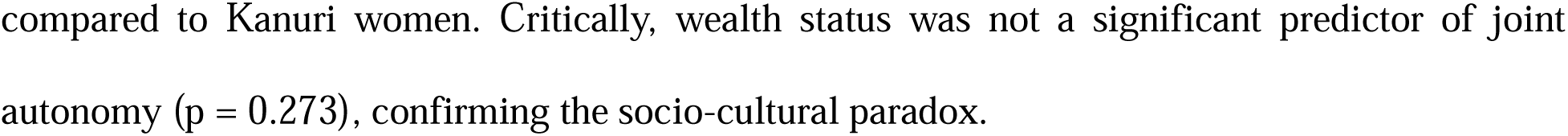
Complex Samples Multinomial Logistic Regression Predicting Healthcare Autonomy Correlates.

Model fit statistics were Cox and Snell 0.047, Nagelkerke 0.062, and McFadden 0.034.

## DISCUSSION

This study assessed the robustness of ethnic and sociocultural determinants of healthcare decision-making autonomy among Hausa, Fulani, and Kanuri women in Northern Nigeria, using complex sample methods to account for the stratified cluster design of the 2024 NDHS. The central finding is that ethnicity and residence are robust determinants of autonomy, persisting after full adjustment for education, wealth, and age. This robustness — the stability of associations across multiple model specifications and covariates — is the principal contribution of the present study and distinguishes it from descriptive cross-tabulations that do not test whether observed differences hold under statistical control. Overall autonomy was severely limited: 72.6% of women reported husband or partner alone decision making, and only 4.9% exercised independent agency. Joint decision making was more common than independent decision making, suggesting that negotiated agency within the household represents a more attainable pathway for female participation than complete independence — a pattern consistent with frameworks that conceptualize empowerment as multidimensional and context-dependent rather than binary [4,7].

Residence was a robust and statistically significant correlate of autonomy (adjusted F = 50.892, p < 0.001). Urban women were substantially more likely to report joint decision making (33.3%) compared with rural women (18.3%), and rural residence was associated with lower adjusted odds of both independent and joint decision making relative to husband-only control. This urban advantage in negotiated agency is consistent with evidence from sub-Saharan Africa suggesting that urban contexts facilitate exposure to health information, broader social networks, and potentially more egalitarian household structures that create space for women’s participation in care-seeking decisions [1]. However, it is important to note that even among urban women in this sample, husband or partner alone decision making remained the majority experience (60.4%), indicating that residential context modifies but does not eliminate the underlying pattern of male-dominated healthcare authority. The rural-urban gradient in autonomy may also partly reflect self-selection processes, whereby women with higher agency are more likely to reside in or migrate to urban areas, which would amplify observed differences without fully representing the causal effect of urbanization itself.

Ethnicity was the most theoretically significant robust correlate identified in this study. After adjustment for all covariates, Hausa and Fulani women had over 60% lower odds of joint decision making compared with Kanuri women (OR: 0.36 and 0.34, respectively), confirming that ethnic identity captures dimensions of household power that conventional socioeconomic indicators do not. This finding is compatible with broader frameworks emphasizing the primacy of gender norms and relational power structures in shaping health behaviours and service uptake [3]. The particularly notable observation is the distinctiveness of the Kanuri/Beriberi pattern: with approximately 41% of Kanuri women reporting joint decision making — nearly double the proportion observed among Hausa and Fulani women — this group exhibits a qualitatively different household dynamic. This difference may reflect the historically distinct social organisation of Kanuri communities in the Lake Chad basin, where certain traditions of spousal consultation in household matters have been documented, or it may reflect differential geographic concentration in Borno State, where local normative environments differ from the broader Northern Nigerian context [5]. Although the Kanuri subsample was considerably smaller (n = 564) than the Hausa (n = 7,087) and Fulani (n = 2,347) subgroups, the magnitude and statistical significance of the ethnic contrasts suggest a genuine socio-cultural differential warranting further qualitative and community-level investigation. From a programmatic standpoint, this ethnic heterogeneity implies that region-wide or ethnicity-blind interventions may be insufficient: strategies to improve women’s healthcare agency must be calibrated to the specific normative landscape of each community.

Education was a statistically significant model effect (Wald F = 3.930, p = 0.001), but its influence followed a gradient rather than a uniform pattern across categories. Women with no formal education had 40% lower odds of joint decision making compared with those with higher education (OR: 0.60), and primary education conferred only marginally better odds (OR: 0.73). Secondary education, however, was not significantly associated with joint decision making (OR: 0.88, CI: 0.71–1.11), suggesting a threshold effect whereby meaningful gains in autonomy may require substantial rather than minimal educational attainment. This is consistent with research indicating that lower levels of schooling may be insufficient to challenge entrenched gender norms in highly traditional settings, and that the relationship between education and autonomy is non-linear in contexts where social structures powerfully constrain individual agency [3,4]. Crucially, wealth status was not a statistically significant predictor after adjustment (Wald F = 1.303, p = 0.273), providing evidence for what this study terms the socio-cultural paradox: the finding that economic resources alone do not confer decision-making authority when gender norms systematically subordinate women’s voice within the household. This paradox has important policy implications, as it suggests that income-generation and poverty-reduction programmes — while valuable in their own right — are unlikely to produce commensurate gains in women’s healthcare autonomy in Northern Nigeria without complementary interventions targeting normative change.

Age was positively and significantly associated with both joint and independent decision making (OR: 1.01 and 1.05, respectively; p < 0.001). Although the per-year effect size is modest, the direction of association is consistent with West African evidence suggesting that older women accumulate social capital within the household over the course of marriage, often through childbearing and extended conjugal tenure, which gradually enhances their bargaining position in domestic decision-making [4]. This implies that interventions targeting younger married women — who face the steepest deficit in healthcare agency — may be particularly important in efforts to reduce maternal and child health inequities in Northern Nigeria.

Several limitations should be considered when interpreting these findings. The cross-sectional design precludes causal inference; observed associations between ethnicity, residence, and autonomy reflect co-occurrence rather than established directional relationships. Healthcare autonomy was operationalized using a single DHS survey item, which, while standard in large-scale demographic research, may not adequately capture the complexity and situational variability of household decision-making processes in practice. Social desirability bias may additionally lead women — particularly in rural and highly patriarchal settings — to under-report their own decision-making participation. Although complex sample procedures were applied throughout, the modest pseudo-R-square values (Nagelkerke R² = 0.062) indicate that a substantial proportion of variation in autonomy remains unexplained, likely attributable to unmeasured factors such as household composition, spousal communication quality, and local community-level normative environments. Future research employing longitudinal designs, qualitative methods, or multilevel modelling with community-level variables would considerably strengthen causal understanding of the pathways through which ethnicity and residence shape women’s healthcare autonomy in Northern Nigeria.

## CONCLUSIONS

Healthcare decision-making autonomy among Hausa, Fulani, and Kanuri women in Northern Nigeria is severely constrained, with husband or partner alone decision-making predominating at 72.6% and fully independent female agency reported by fewer than 5% of the sample. The central finding of this study is that ethnicity and residential context are robust determinants of autonomy, persisting independently after adjustment for education, wealth, and age. This robustness confirms that sociocultural and structural factors — tied to ethnic identity and geographic context — exert an influence on women’s healthcare agency that conventional socioeconomic interventions alone cannot address. The non-significance of wealth status after full adjustment provides compelling empirical support for the socio-cultural paradox: that economic resources do not translate into decision-making authority when gender norms systematically subordinate women’s voice within the household. These findings carry direct and specific recommendations for policymakers and health programme designers. First, interventions targeting women’s healthcare autonomy in Northern Nigeria must be ethnicity-sensitive and community-specific. The markedly different autonomy profiles observed across Hausa, Fulani, and Kanuri communities indicate that blanket regional approaches are insufficient. Programmes should be co-designed with community leaders, religious figures, and women’s groups within each ethnic community, addressing the particular household norms and power structures that govern decision-making in that context. Second, male engagement must be an explicit and structured component of any autonomy-strengthening intervention. Since husbands and partners function as primary gatekeepers of healthcare access for the majority of women in this sample, interventions that focus exclusively on women are unlikely to shift household dynamics. Community health worker programmes, spousal communication workshops, and gender-transformative dialogue initiatives should be prioritised, particularly in rural settings where the autonomy deficit is most severe. Third, rural communities must be designated as the highest-priority settings for such interventions. The 15-percentage-point gap in joint decision-making between rural (18.3%) and urban (33.3%) women, sustained after adjustment for all covariates, indicates that rural residence captures structural disadvantages — including geographic isolation, limited exposure to health information, and stronger enforcement of traditional gender norms — that require dedicated policy attention. Fourth, while education programmes remain important, the threshold effect observed in this study suggests that investment must target higher levels of attainment. Primary education alone is insufficient to meaningfully shift autonomy outcomes; sustained investment in secondary and higher education for girls and women is necessary to achieve the levels of educational attainment that appear to confer meaningful gains in healthcare agency. Future research should build on the present findings in several directions. Longitudinal studies are needed to establish whether the observed associations between ethnicity, residence, and autonomy reflect causal pathways or confounded cross-sectional co-occurrence. Qualitative and mixed-methods research is particularly warranted to explore the mechanisms through which ethnic identity shapes household decision-making dynamics — including the specific norms, communication patterns, and community enforcement processes that differ across Hausa, Fulani, and Kanuri communities. Multilevel modelling incorporating community-level variables such as local gender norm indices, community health infrastructure, and female leadership representation would allow researchers to disentangle individual from contextual determinants of autonomy more precisely. Additionally, the distinctively higher joint decision-making rates observed among Kanuri women deserve dedicated investigation; understanding what cultural, historical, or structural features of Kanuri communities support more collaborative household dynamics could offer important lessons for norm-change interventions in Hausa and Fulani communities. Finally, future studies should examine autonomy across additional healthcare decision types — including contraceptive use, child healthcare, and antenatal care — to determine whether the ethnic and residential patterns identified here generalise across the full spectrum of women’s health decision-making in Northern Nigeria.

## DECLARATIONS

### Ethics approval and consent to participate

This study is a secondary analysis of anonymized NDHS 2024 data obtained with permission from the DHS Program. The NDHS protocols are implemented with ethical approval and with informed consent obtained from participants prior to interview. For more information on the ethical standards of DHS data, please visit:https://dhsprogram.com/Methodology/Protecting-the-Privacy-of-DHS-Survey-Respondents.cfm.

### Consent for Publication

Not applicable

### Availability of data and materials

The NDHS datasets are available from the DHS Program upon reasonable request/registration. These data remain downloadable, contingent on the website’s active status: https://dhsprogram.com/data/dataset/Nigeria_Standard-DHS_2024.cfm?flag=0

## Funding

No specific funding was received for this study.

### Authors’ contributions

Abolaji Moses OGUNETIMOJU: Conceptualization, study design, methodology oversight, investigation, writing – original draft (lead), writing – review and editing (lead), visualization oversight, supervision, project administration. Samuel Ayoola AJEBORIOGBON: Data curation, formal analysis (primary), software, validation, writing – original draft (support), writing – review and editing (support), visualization (primary). Both authors read and approved the final manuscript, and agree to be accountable for all aspects of the work.

## Data Availability

All data produced are available online at https://dhsprogram.com/data/dataset/Nigeria_Standard-DHS_2024.cfm?flag=0

## Acknowledgements

We express our profound gratitude to the women across Northern Nigeria who participated in the 2024 Nigeria Demographic and Health Survey. Their openness in sharing their lived experiences provides the essential foundation for understanding the complexities of gender equity and healthcare agency in the region.

